# Endovascular thrombectomy for mild vertebrobasilar artery occlusion: a hierarchical win-ratio analysis

**DOI:** 10.64898/2026.09.18.26363452

**Authors:** Yapeng Guo, Xinyu Fan, Yingjie Xu, Xinru Zhou, Wei Li, Junfeng Xu, Zhixin Huang, Wensheng Zhang, Xianjun Huang

**Affiliations:** Department of Neurology, The First Affiliated Hospital of Wannan Medical University (Yijishan Hospital of Wannan Medical University), Wuhu, Anhui Province, China; Department of Neurology, Centre for Leading Medicine and Advanced Technologies of IHM, The First Affiliated Hospital of USTC, Division of Life Sciences and Medicine, University of Science and Technology of China, Hefei, Anhui, China; Department of Neurology, The First Affiliated Hospital of Hainan Medical University, Haikou, Hainan Province, China; Department of Neurology, The Affiliated Guangdong Second Provincial General Hospital of Jinan University, Guangzhou, China; Department of Neurology, Heyuan People’s Hospital, Heyuan, Guangdong Province, China

**Keywords:** vertebrobasilar artery occlusion, endovascular thrombectomy, mild stroke, NIHSS, win statistics, death status

## Abstract

**Background:** Endovascular thrombectomy (EVT) is used selectively for vertebrobasilar artery occlusion (VBAO), but its net value in mild deficits remains uncertain. We performed a hierarchical win-ratio analysis of EVT versus best medical management (BMM) in patients with acute VBAO and admission National Institutes of Health Stroke Scale (NIHSS) scores of 10 or less.

**Methods:** We performed a secondary analysis of a multicenter retrospective registry of acute VBAO with admission NIHSS scores of 10 or less. EVT plus BMM was compared with BMM alone in the overall cohort and in admission NIHSS strata of 0-5 and 6-10. The primary hierarchy ranked death status through common follow-up, 90-day modified Rankin Scale score, and symptomatic intracranial hemorrhage (sICH). All EVT-BMM pairs were compared sequentially and classified as an EVT win, BMM win, or tie. The adjusted analysis used stabilized inverse probability of treatment weighting (IPTW), with weights truncated at the 1st and 99th percentiles in a sensitivity analysis.

**Results:** The analysis included 1,232 patients: 428 underwent EVT and 804 received BMM. In the unweighted overall analysis, EVT had 146,674 wins, BMM had 149,958 wins, and 47,480 ties, yielding a WR of 0.98 (95% CI, 0.84-1.15; P = 0.749). After stabilized IPTW, the WR was 0.96 (95% CI, 0.80-1.14; P = 0.618). Sensitivity analyses did not show a robust overall association favoring EVT. In exploratory NIHSS-stratified analyses, the WR was 0.80 (95% CI, 0.61-1.05; P = 0.101) for NIHSS scores of 0-5 and 1.36 (95% CI, 1.10-1.68; P = 0.004) for scores of 6-10.

**Conclusions:** EVT was not associated with an overall net advantage over BMM in this hierarchical analysis. The exploratory signal among patients with admission NIHSS scores of 6-10 suggests that EVT may be feasible in selected patients and warrants confirmation in randomized trials; for patients with NIHSS scores of 0-5, EVT use should remain cautious and individualized.

**Study registration:** ChiCTR2000033211, www.chictr.org.cn/showproj.html?proj=54046

**Clinical Perspective:** *What Is New?:*

- In this secondary analysis of 1,232 patients with acute vertebrobasilar artery occlusion and admission NIHSS scores of 10 or less, a hierarchical outcome prioritizing death status, 90-day disability, and symptomatic intracranial hemorrhage did not show an overall net advantage of endovascular thrombectomy plus best medical management over best medical management alone.
- Exploratory analyses suggested heterogeneity within the mild-deficit range: the win ratio was 0.80 (95% CI, 0.61-1.05) for NIHSS scores of 0-5 and 1.36 (95% CI, 1.10-1.68) for scores of 6-10. These findings should not be interpreted as a treatment threshold.

*What Are the Clinical Implications?:*

- Mild vertebrobasilar artery occlusion should not be considered a uniform indication for routine thrombectomy or routine conservative management.
- Treatment selection should integrate neurological severity with treatment timing, infarct burden, occlusion mechanism and location, collateral or perfusion status, and anticipated procedural risk and benefit. Prospective studies are needed to validate these findings.

## INTRODUCTION

Acute vertebrobasilar artery occlusion (VBAO) is a high-risk posterior circulation stroke that can cause early neurological deterioration, severe disability, or death. Endovascular thrombectomy (EVT) has shown benefit in selected patients with VBAO, especially those with moderate-to-severe deficits.^1,2^ However, its value in patients with low National Institutes of Health Stroke Scale (NIHSS) scores remains uncertain.^3–5^

In posterior – circulation stroke, a low NIHSS score may underestimate the clinical risk posed by a severe vascular lesion. Several studies have therefore examined whether EVT improves outcomes in patients with low-NIHSS VBAO stroke, but their findings remain inconsistent.^6–8^ One recent study reported that EVT did not improve functional prognosis and was associated with increased mortality.^7^ In contrast, another study suggested that EVT improved functional outcomes, although this benefit was accompanied by a higher rate of symptomatic intracranial hemorrhage (sICH).^9^ However, investigators have questioned whether the modified Rankin Scale (mRS) alone is sufficient as a primary endpoint for neurointerventional trials and have advocated broader use of complementary outcome measures, particularly in patients with low-NIHSS stroke.^10^

A hierarchical win-ratio framework can address part of this problem by comparing patients pairwise across treatment groups using a prespecified hierarchy.^11^ Each pair is first compared on the most clinically important outcome; if tied, the comparison proceeds to the next component.^12^ This approach can prioritize death status, functional status, and safety while preserving component-level interpretation.

We therefore performed a hierarchical win-ratio reanalysis of EVT versus BMM using the registry data. The aim was to quantify the net direction of outcome differences and safety signals when death status, 90-day disability, and sICH were integrated into a prioritized outcome. We also examined whether the hierarchical result differed between NIHSS scores of 5 or less and greater than 5, because low-deficit VBAO may be clinically heterogeneous.

## METHODS

### Study Population

This retrospective multicenter cohort study used data from the registry, which included patients with acute VBAO treated at 65 stroke centers in 15 provinces in China between December 2015 and June 2022.^13^ Eligible patients were 18 years or older, had VBAO confirmed by CT angiography, MR angiography, or digital subtraction angiography (DSA), presented within 24 hours of last known well, and had an admission NIHSS score of 10 or less.

Among 1,427 eligible patients with VBAO and admission NIHSS scores of 10 or less, 62 (4.3%) had missing 90-day mRS data and 133 (9.3%) had no ascertainable 1-year mortality status; these exclusions accounted for the 195 patients not included in the analytic cohort. The final complete-case analytic cohort included 1,232 patients. No imputation was performed, and no missing values were present for variables used in the win-ratio or inverse probability of treatment weighting (IPTW) models; door-to-puncture time was structurally unavailable for patients who did not undergo EVT. Patients were classified as receiving EVT plus BMM or BMM alone according to the treatment actually received (Figure 1).

**Figure 1.**
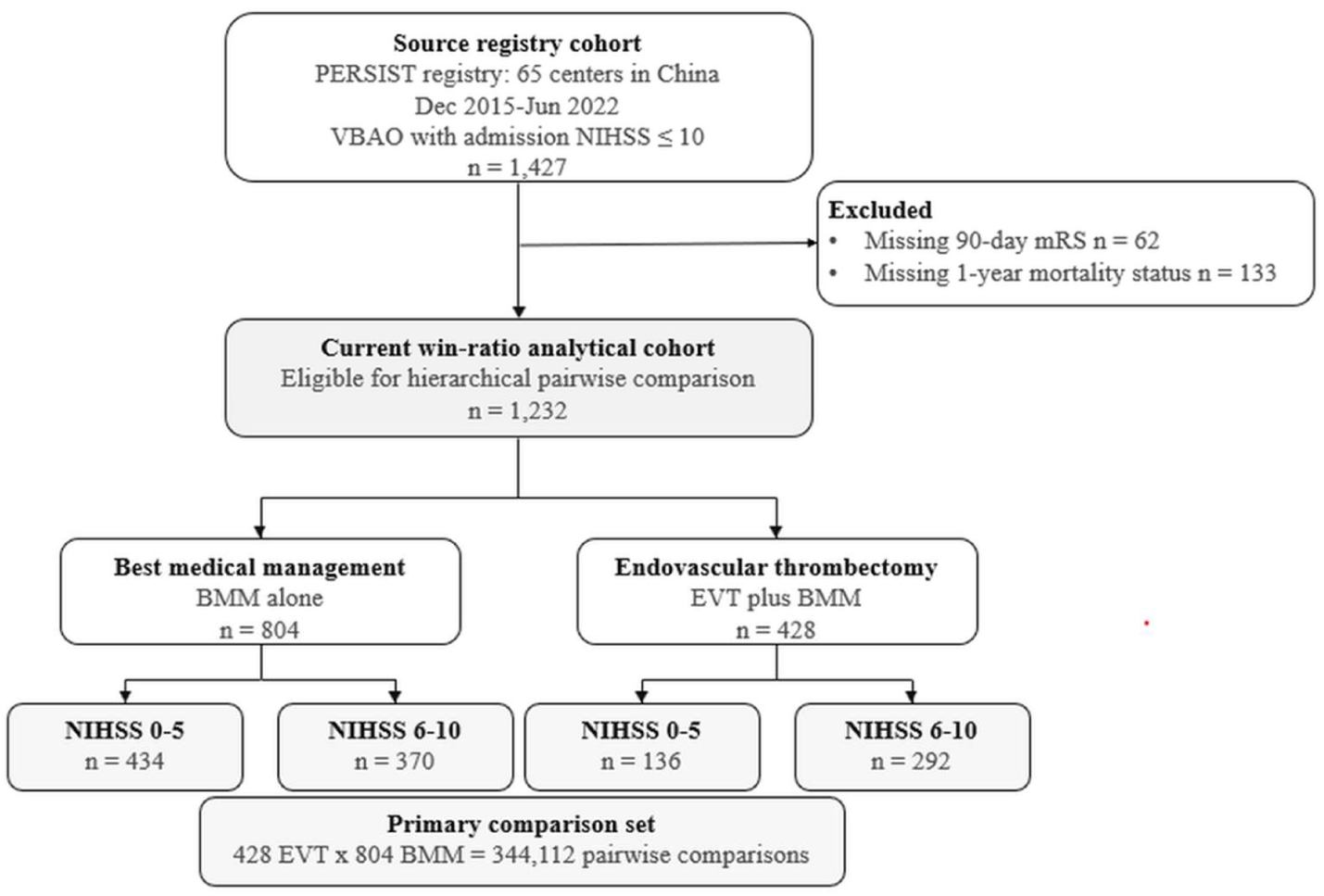
Patient flowchart. BMM, best medical management; EVT, endovascular thrombectomy; mRS, modified Rankin scale; NIHSS, National Institutes of Health Stroke Scale; PERSIST, PostErior ciRculation iSchemIc Stroke registry; VBAO, vertebrobasilar artery occlusion.

The registry was approved by the ethics committee of the First Affiliated Hospital of the University of Science and Technology of China and participating centers (no. 2020–40). The requirement for informed consent was waived because of the retrospective study design.

### Data Collection

Demographic, clinical, vascular risk factor, imaging, treatment, time-metric, and follow-up data were collected by retrospective review of medical records. Baseline variables included age, sex, vascular risk factors, prestroke mRS, admission NIHSS score, posterior circulation Alberta Stroke Prognosis Early CT Score (pc-ASPECTS), occlusion site, Trial of Org 10172 in Acute Stroke Treatment (TOAST) subtype,^14^ intravenous thrombolysis (IVT), blood pressure, and last-seen-well interval. Stroke severity was assessed with the NIHSS. pc-ASPECTS was evaluated on noncontrast CT or MRI. BMM denoted BMM alone, whereas EVT denoted EVT in addition to BMM. Door-to-puncture time was defined as the interval from hospital arrival to arterial puncture and was recorded only for EVT-treated patients

Follow-up outcomes included mRS scores at 90 days and 1 year, mortality at 90 days and 1 year, and sICH. sICH was defined according to the Heidelberg Bleeding Classification as newly observed intracranial hemorrhage on imaging associated with either an increase of at least 4 points in the total NIHSS score or more than 2 points in one NIHSS item, or clinical deterioration requiring hemicraniectomy, intubation, external ventricular drainage, or another major medical intervention.^15^

### Outcome Hierarchy

The outcome hierarchy was prespecified after discussion among stroke neurologists, neurointerventionalists, and statisticians, following published win-ratio recommendations that components should be ordered by clinical importance. Consensus was reached to rank death first, followed by 90-day mRS and then sICH, because survival has the highest patient-centered priority, functional disability is the standard efficacy outcome in stroke trials, and sICH represents an important safety event that is most clinically interpretable after mortality and disability have been considered. Mortality was assessed at two follow-up time points: 90 days and 1 year after stroke onset. For each EVT-BMM patient pair, only deaths occurring within the follow-up period shared by both patients were considered. If one patient died within the shared follow-up period and the other remained alive, the patient who remained alive was classified as the winner. If both patients died in different prespecified intervals, the patient with the longer survival interval was classified as the winner. Operationally, death within 90 days was ranked worse than death after 90 days and within 1 year, and both were ranked worse than being alive at 1 year. If the available mortality follow-up did not distinguish the pair, the pair was tied on death and moved to the next hierarchy level.

If a pair was tied on death, the comparison proceeded to 90-day mRS score, with lower scores considered better. If the pair remained tied, sICH was compared, with absence of sICH considered better.

### Descriptive and Conventional Statistical Analysis

Continuous variables were summarized as medians with interquartile ranges and compared using the Mann–Whitney U test. Categorical variables were summarized as counts and percentages and compared using the chi-square test or Fisher exact test, as appropriate. Covariate balance before and after IPTW was assessed using absolute standardized mean differences, with values below 0.10 indicating adequate balance. The analytic cohort was complete for all variables used in the win-ratio and IPTW models, so no imputation was performed. Door-to-puncture time was missing by design in the BMM group and was not used for propensity-score estimation.

### Win-Ratio Analysis

All possible pairwise comparisons were generated between EVT-treated and BMM-treated patients. Each pair was compared sequentially according to the prespecified hierarchy. A pair was counted as an EVT win if the EVT-treated patient had the better outcome at the first component that separated the pair, and as a BMM win if the BMM-treated patient had the better outcome. Pairs tied across all components were classified as ties.

The win ratio (WR) was calculated as the number of EVT wins divided by the number of BMM wins. A WR greater than 1 favored EVT, and a WR less than 1 favored BMM. Win odds were calculated as (EVT wins + 0.5 × ties) / (BMM wins + 0.5 × ties). Confidence intervals (CI) and P values for the primary, NIHSS-stratified and truncated-IPTW win-ratio analyses were estimated using 1,000 bootstrap resamples. Win statistics were implemented in R and cross-checked using the BuyseTest package.^16,17^

### IPTW

Because treatment was not randomly assigned, stabilized IPTW was used for the adjusted overall EVT-versus-BMM analysis. Propensity scores for receiving EVT were estimated with logistic regression using age, sex, diabetes, prior stroke or transient ischemic attack, admission NIHSS score, pc-ASPECTS, TOAST subtype, IVT, prestroke mRS, occlusion site, systolic blood pressure, hypertension, last-seen-well interval greater than 6 hours, hyperlipidemia, coronary heart disease, atrial fibrillation, diastolic blood pressure, and smoking. Door-to-puncture time was not included because it was undefined for patients who did not undergo EVT. Stabilized weights were incorporated into the pairwise win-counting procedure. The observed stabilized weights ranged from 0.39 to 5.69. A sensitivity analysis truncated weights at the 1st and 99th percentiles (0.41 and 3.60, respectively).

For the IPTW win-ratio analysis, each EVT-BMM pair contributed according to the product of the stabilized weights of the two patients; wins, losses, ties, and total pair counts are therefore reported as weighted pair counts.

### Subgroup and Sensitivity Analyses

Subgroup analyses were performed according to admission NIHSS scores of 0-5 versus 6-10. The interaction P value was obtained by bootstrap comparison of log win ratios between the two prespecified NIHSS strata and was interpreted as exploratory rather than as definitive evidence of treatment-effect modification. Sensitivity analyses tested alternative endpoint orders. Model 1 ranked 90-day mRS, death status through common follow-up, and sICH. Model 2 ranked death status through common follow-up, sICH, and 90-day mRS. Model 3 ranked 90-day mRS, sICH, and death status through common follow-up. Model 4 included death status through common follow-up and 90-day mRS only, omitting sICH.

### Statistical Software

All analyses were performed in R 4.6.0. Win statistics were implemented with reproducible R scripts and validated using BuyseTest. IPTW, descriptive analyses, and figure generation were performed using R packages. Two-sided P values less than 0.05 were considered statistically significant.

## RESULTS

### Study Population

The analysis included 1,232 patients with acute VBAO and mild neurological deficits, of whom 804 (65.3%) received BMM and 428 (34.7%) underwent EVT. Before weighting, EVT-treated patients were younger than BMM-treated patients (median age, 65.0 [55.0-72.0] vs 66.0 [58.0-75.0] years; P < 0.001) and had higher admission NIHSS scores (7 [4-9] vs 5 [3-7]; P < 0.001). EVT-treated patients were less likely to receive IVT (18.7% vs 26.0%; P = 0.004), had a higher proportion of large-artery atherosclerosis (TOAST distribution, P < 0.001), and differed in occlusion-site distribution (P < 0.001). The proportion last seen well more than 6 hours previously was similar between groups (41.1% vs 36.9%; P = 0.151). Baseline characteristics within the NIHSS strata are shown in Table 1, and overall unweighted characteristics are provided in Table S1. After stabilized IPTW, all measured covariates included in the propensity-score model were balanced, with a maximum absolute standardized mean difference of 0.067 (Figure S1).

**Table 1.**
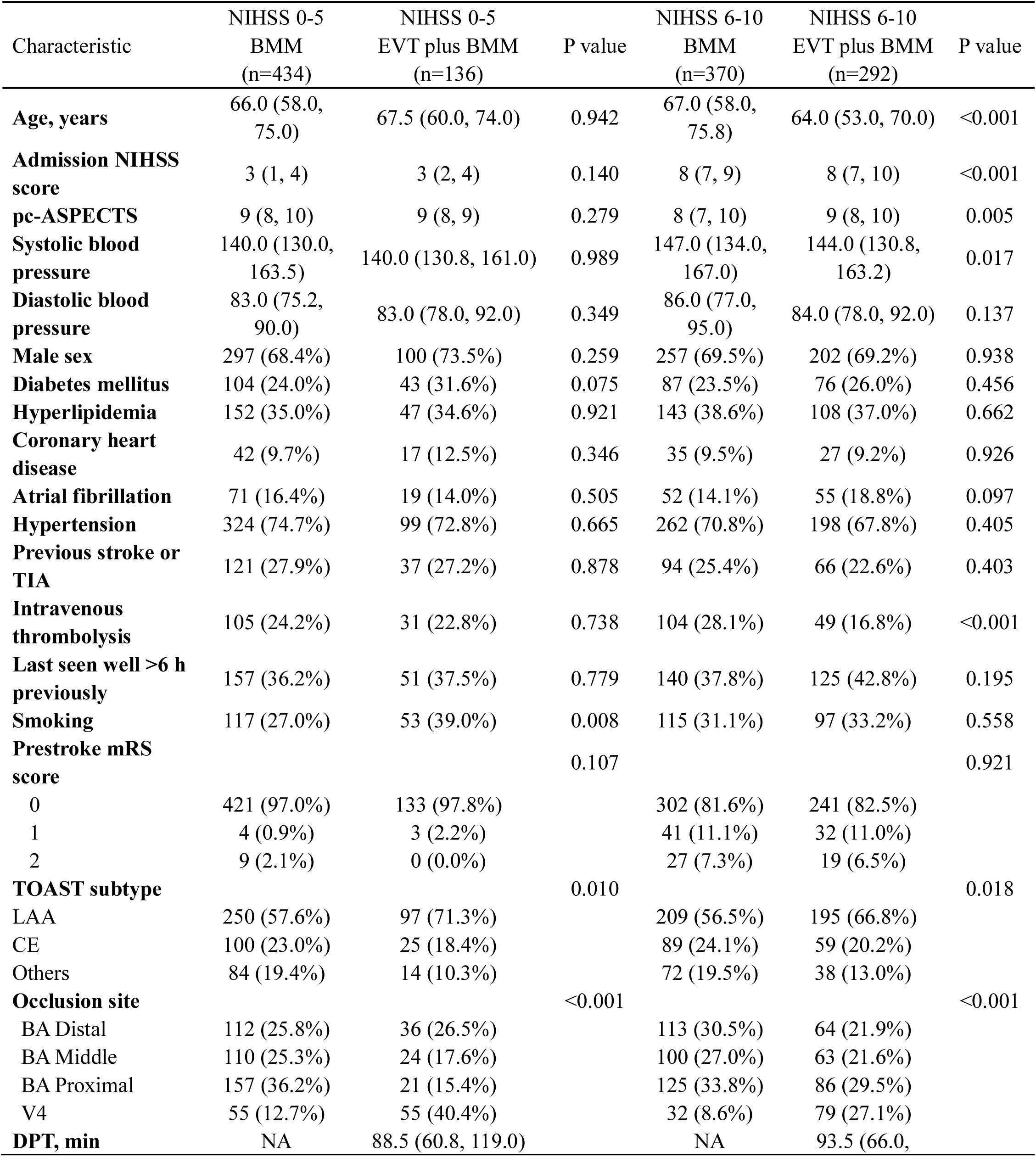
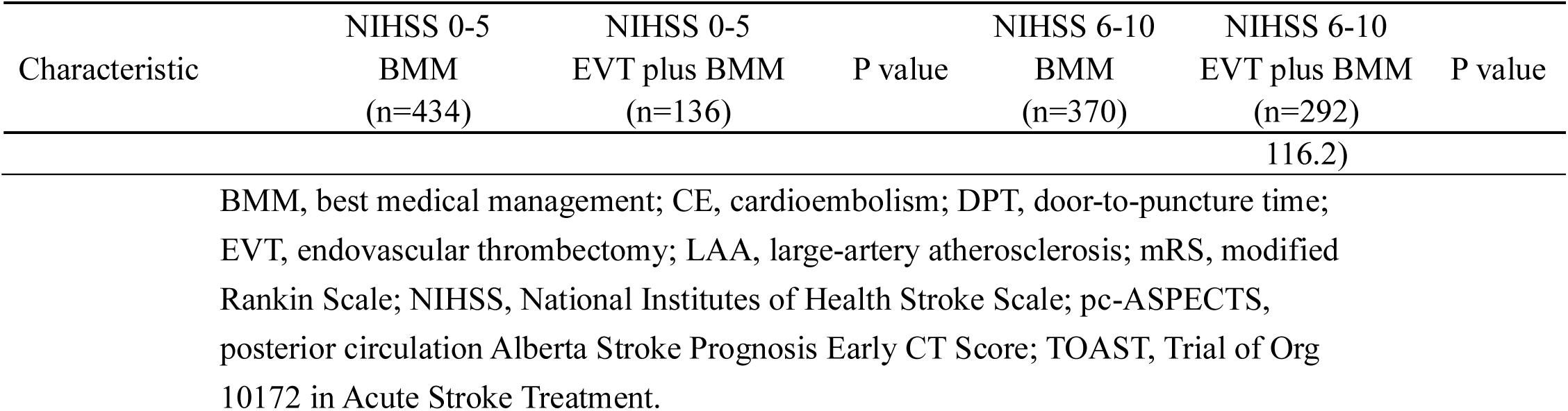
Baseline characteristics by treatment group within admission NIHSS strata.

### Observed Outcomes

The median 90-day mRS score was 2 in both groups, with an interquartile range (IQR) of 1-4 in the BMM group and 1-4 in the EVT group. Ninety-day mortality occurred in 92 of 804 (11.4%) BMM-treated patients and 58 of 428 (13.6%) EVT-treated patients. One-year mortality occurred in 110 (13.7%) BMM-treated patients and 70 (16.4%) EVT-treated patients. Death after 90 days and within 1 year was observed in 18 (2.2%) BMM-treated patients and 12 (2.8%) EVT-treated patients. sICH occurred in 17 (2.1%) BMM-treated patients and 30 (7.0%) EVT-treated patients (Table 2).

**Table 2.** Outcomes by treatment group in total cohort.

| Treatment | No. of patients | 90-day death | 1-year death | 90-day mRS, median [IQR] | sICH |
| --- | --- | --- | --- | --- | --- |
| BMM | 804 | 92 (11.4%) | 110 (13.7%) | 2 [1, 4] | 17 (2.1%) |
| EVT plus BMM | 428 | 58 (13.6%) | 70 (16.4%) | 2 [1, 4] | 30 (7.0%) |

BMM, best medical management; EVT, endovascular thrombectomy; IQR, interquartile range; mRS, modified Rankin Scale; sICH, symptomatic intracranial hemorrhage.

### Overall Win-Ratio Analysis

In the unweighted overall analysis, 428 EVT-treated patients were compared with 804 BMM-treated patients, producing 344,112 pairwise comparisons. EVT won 146,674 comparisons (42.6%), BMM won 149,958 comparisons (43.6%), and 47,480 comparisons (13.8%) remained tied. The resulting WR was 0.98 (95% CI, 0.84-1.15; P = 0.749), and the WO was 0.98 (Figure 2).

**Figure 2.**
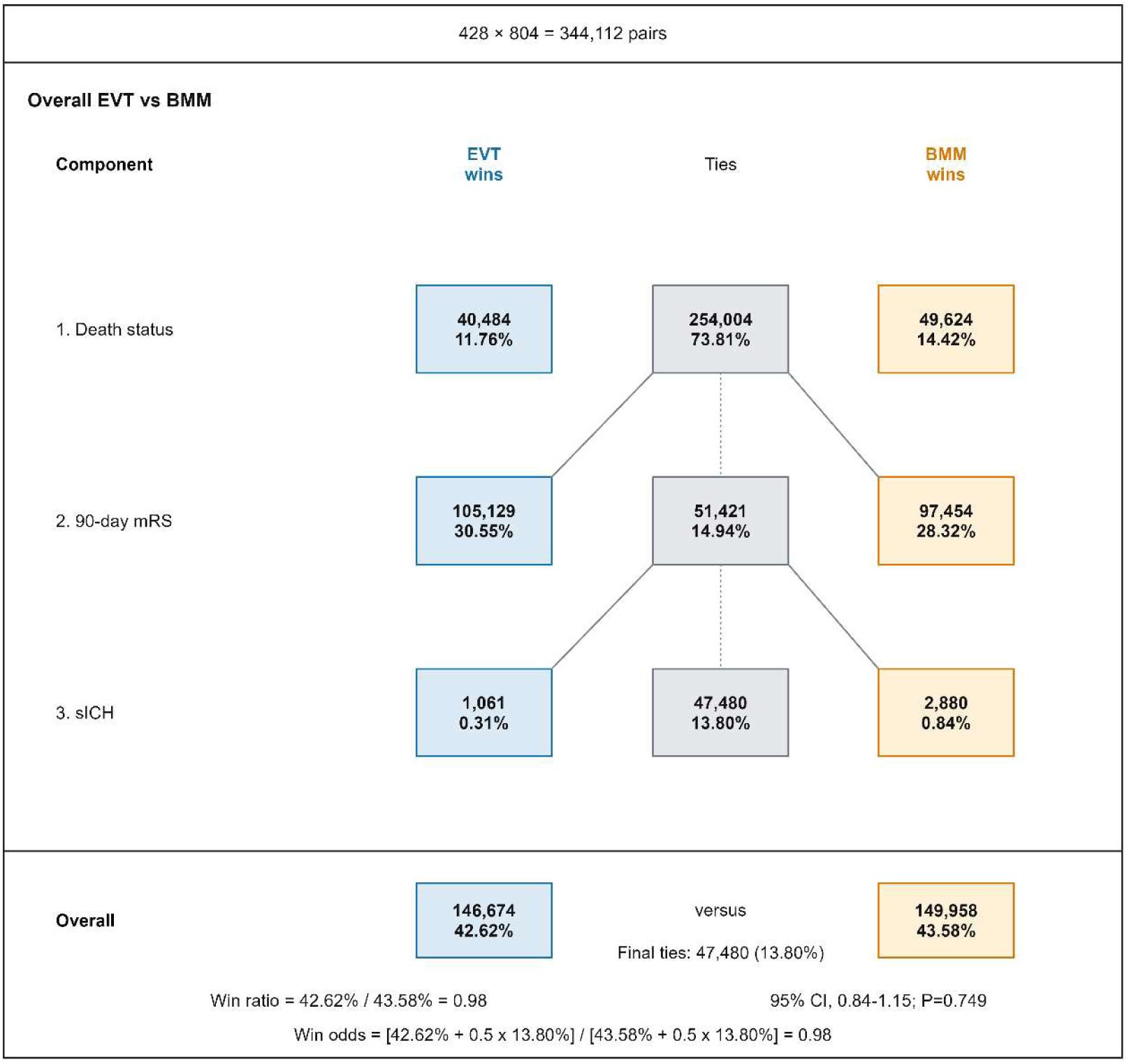
Overall win-ratio flow diagram for EVT versus BMM. BMM, best medical management; CI, confidence interval; EVT, endovascular thrombectomy; mRS, modified Rankin Scale; sICH, symptomatic intracranial hemorrhage; WO, win odds; WR, win ratio.

The component-level analysis showed the distribution of pairwise wins across the hierarchy. At the death-status component, EVT won 40,484 comparisons (11.8%) and BMM won 49,624 comparisons (14.4%), with 254,004 comparisons (73.8%) moving forward as ties. At the 90-day mRS component, EVT won 105,129 comparisons (30.6%) and BMM won 97,454 comparisons (28.3%). At the sICH component, EVT won 1,061 comparisons (0.3%) and BMM won 2,880 comparisons (0.8%). Component-level win counts are shown in Figure 2.

### IPTW-Adjusted Overall Analysis

In the stabilized IPTW analysis, the weighted pair total was 346,456.2. EVT won 146,720.9 weighted comparisons (42.35%), BMM won 153,358.3 weighted comparisons (44.26%), and 46,377.1 weighted comparisons (13.39%) remained tied. The weighted WR was 0.96 (95% CI, 0.80-1.14; P = 0.618), and the weighted WO was 0.96. At the death-status component, EVT and BMM contributed 41,356.4 and 49,975.8 weighted wins, respectively. The corresponding weighted wins were 104,345.3 and 99,454.1 at the 90-day mRS component and 1,019.2 and 3,928.4 at the sICH component (Figure S2).

The weighted pair counts are non-integer because each pair contributes according to the product of the stabilized treatment weights (Table 3).

**Table 3.**
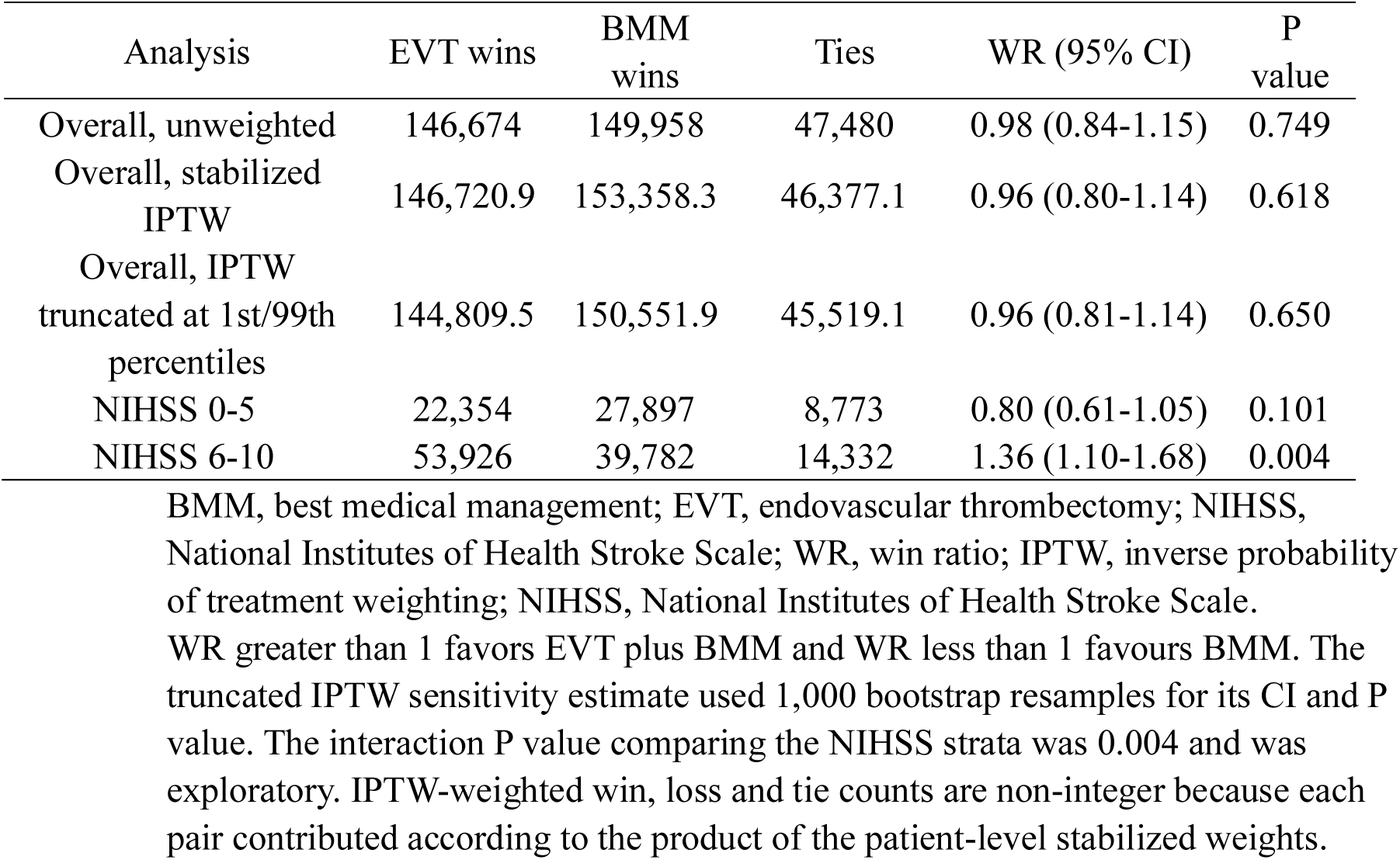
Overall and NIHSS-stratified hierarchical win-ratio analyses.

### Sensitivity Analyses

Across the four prespecified alternative endpoint hierarchies, the WR estimates ranged from 0.96 to 0.99 and remained close to unity; the direction of the overall estimate did not materially change (Figure S3). Truncating the stabilized IPTW weights at the 1st and 99th percentiles yielded a WR of 0.96 (95% CI, 0.81-1.14; P = 0.650; Table 3).

### NIHSS-Stratified Analyses

The NIHSS-stratified results suggested heterogeneity across the mild-deficit range. Among patients with admission NIHSS scores of 5 or less, 136 EVT-treated patients were compared with 434 BMM-treated patients, producing 59,024 pairwise comparisons. EVT won 22,354 comparisons (37.9%), BMM won 27,897 comparisons (47.3%), and 8,773 comparisons (14.9%) remained tied. The WR was 0.80 (95% CI, 0.61-1.05; P = 0.101), and the WO was 0.83 (Figure 3).

**Figure 3.**
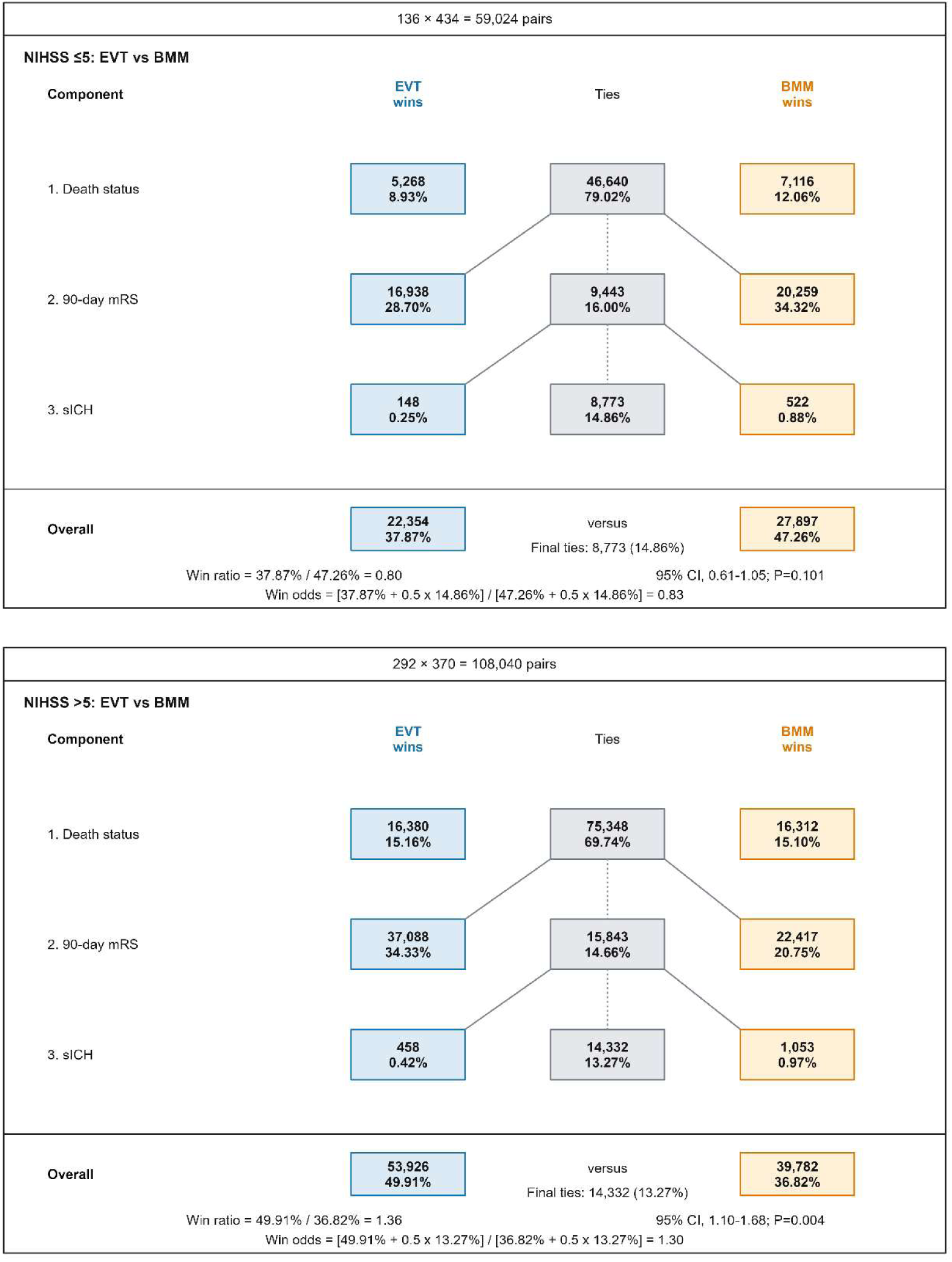
NIHSS-stratified win-ratio flow diagrams. BMM, best medical management; CI, confidence interval; EVT, endovascular thrombectomy; NIHSS, National Institutes of Health Stroke Scale; WR, win ratio.

The unadjusted mRS distributions in the NIHSS 0-5 stratum are shown descriptively in Figure 4. An mRS score of 0-3 was observed in 345 of 434 (79.5%) BMM-treated patients and 97 of 136 (71.3%) EVT-treated patients at 90 days, and in 362 of 434 (83.4%) and 103 of 136 (75.7%), respectively, at 1 year. mRS category 6 (death) accounted for 38 of 434 (8.8%) and 16 of 136 (11.8%) patients at 90 days and 44 of 434 (10.1%) and 18 of 136 (13.2%) at 1 year. These distributions were not adjusted for treatment-selection differences and were not subjected to a separate hypothesis test.

**Figure 4.**
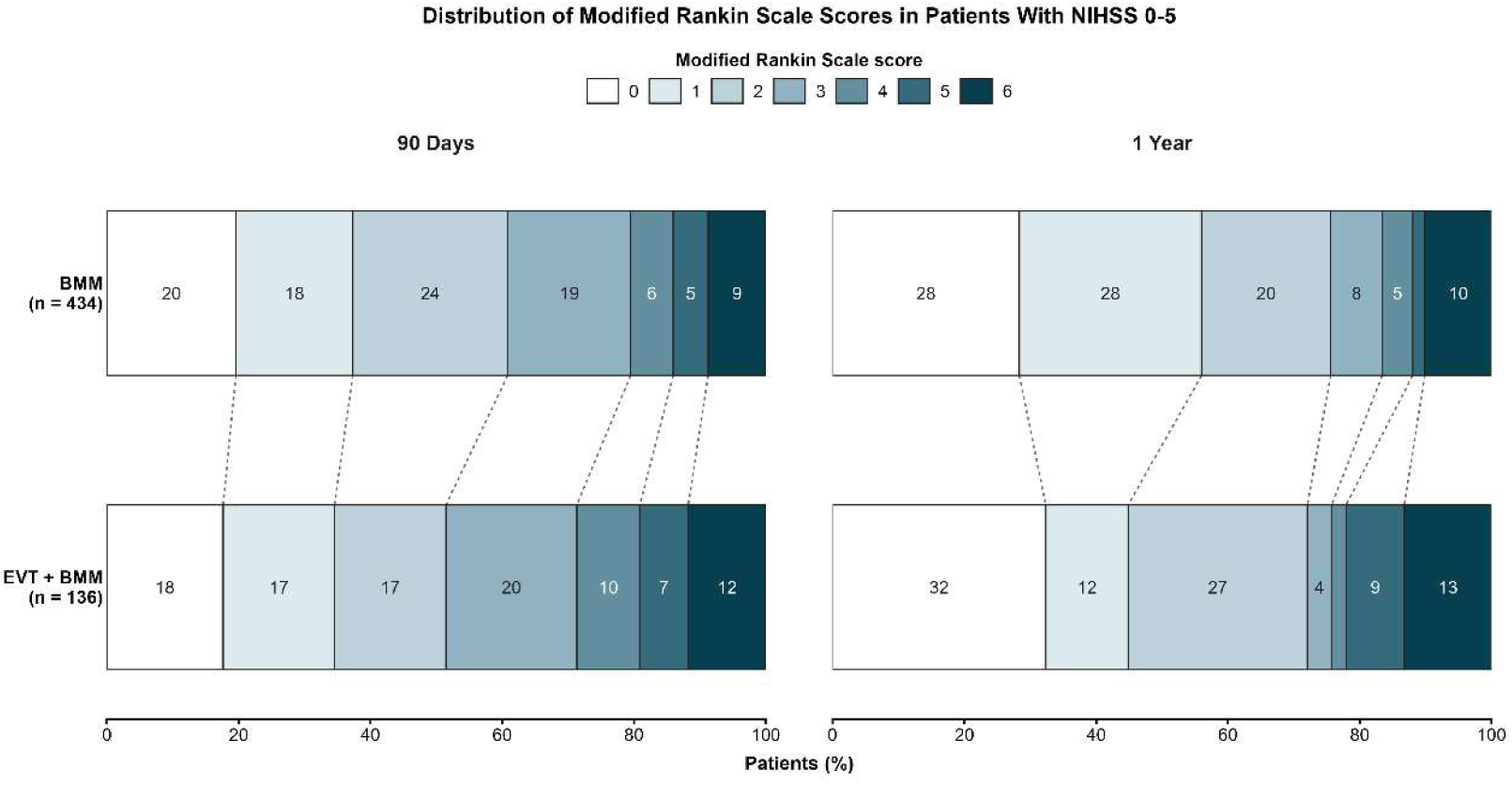
Distribution of modified Rankin Scale scores at 90 days and 1 year among patients with admission NIHSS scores of 0-5. Percentages may not sum to 100% because of rounding. BMM, best medical management; EVT, endovascular thrombectomy; mRS, modified Rankin Scale; NIHSS, National Institutes of Health Stroke Scale.

Among patients with admission NIHSS scores greater than 5, 292 EVT-treated patients were compared with 370 BMM-treated patients, producing 108,040 pairwise comparisons. EVT won 53,926 comparisons (49.9%), BMM won 39,782 comparisons (36.8%), and 14,332 comparisons (13.3%) remained tied. The WR was 1.36 (95% CI, 1.10-1.68; P = 0.004), and the WO was 1.30. In this subgroup, the functional component contributed substantially to the favorable EVT result: EVT won 37,088 comparisons on 90-day mRS, compared with 22,417 BMM wins at that component. The bootstrap comparison of log win ratios yielded an exploratory interaction P value of 0.004; because the subgroup analyses were unadjusted and subject to substantial within-stratum baseline imbalance, this finding should not be interpreted as causal treatment-effect modification (Table 3; Figure 3).

## DISCUSSION

In this hierarchical win-ratio analysis of 1,232 patients with VBAO and mild neurological deficits, EVT was not associated with an overall prioritized hierarchical comparison over BMM when death status, 90-day disability, and sICH were evaluated in clinical priority order. This neutral finding was consistent in the unweighted analysis and after stabilized IPTW. The overall estimate, however, concealed clinically relevant heterogeneity within the low-NIHSS range. EVT was not favored in patients with NIHSS scores of 5 or less, whereas patients with NIHSS scores greater than 5 showed a favorable hierarchical signal (WR, 1.36; 95% CI, 1.10-1.68).

The ATTENTION and BAOCHE trials have established the benefit of EVT in selected patients with VBAO, but their enrolled populations were weighted toward moderate-to-severe deficits and provide limited direct evidence for patients with low NIHSS scores.^3,4^ Recently, several studies investigated the effectiveness of EVT for VBAO patients with low-to-moderate NIHSS presentations; however, the results were controversial.^6,18^ Our analysis provides one explanation for these divergent results by shifting the estimand from a single functional threshold to a prioritized net clinical comparison. In mild-deficit VBAO, a treatment may improve the disability distribution yet fail to produce a net advantage if those gains are counterbalanced by death or sICH.

The component-level results support this explanation. EVT produced more wins at the 90-day mRS component, indicating a potential functional advantage in selected patients.^9^ This signal was offset by fewer favorable comparisons at the death-status component and by more unfavorable comparisons at the sICH component.^18^ The pattern is clinically plausible. Patients with very low NIHSS scores often have limited measurable room for improvement on conventional mRS thresholds, while exposure to arterial puncture, device manipulation, rescue angioplasty or stenting, antithrombotic escalation, anesthesia, and reperfusion-related hemorrhage remain real.^19,20^ In such patients, a modest functional shift may not compensate for even a small excess of serious safety events. Conversely, patients with NIHSS scores above 5 may have more threatened tissue, greater potential for functional recovery, or a higher risk of deterioration without recanalization, making the same procedural risk more acceptable.

The NIHSS-stratified findings reinforce that mild posterior-circulation stroke is not a uniform clinical state. In our cohort, unadjusted 90-day and 1-year mRS distributions showed no descriptive EVT advantage at NIHSS 0-5, consistent with prior observational findings.^19^ Early neurological deterioration has been reported in medically managed VBAO and minor stroke with large-vessel occlusion.^21,22^ However, this dataset did not specifically capture deterioration and therefore cannot determine its frequency or define the role of rescue EVT. In patients with NIHSS scores of 6-10, the favorable exploratory hierarchical signal suggests that selected patients may benefit more from reperfusion. This finding aligns with reports of stronger EVT associations among patients with NIHSS scores of 6-9 than among those with lower scores.^6^ These findings support prospective EVT evaluation in selected patients with scores of 6-10 and cautious, individualized management at scores of 0-5. Overall, treatment decisions should remain individualized rather than rely on NIHSS alone.

Although conventional dichotomized mRS analyses remain commonly used in stroke trials, the win-ratio framework is better suited for patients with mild neurological deficits because it makes the clinical hierarchy explicit.^23^ Hierarchical pairwise comparison permits death status, disability, and sICH to be evaluated within a single estimand while preserving component-level interpretation.^20,24^ In the present study, this approach showed that the functional component did not translate into an overall net advantage once survival and hemorrhagic harm were incorporated. This framework reflects the bedside decision problem in mild VBAO, where clinicians, patients, and families must weigh the possibility of preventing disability against the possibility of treatment-related harm.

The findings should therefore be interpreted as hypothesis-generating rather than practice-changing. The neutral overall WR does not prove that EVT is ineffective in all patients with mild VBAO. It indicates that, when EVT was evaluated as an undifferentiated binary strategy in this registry population, it was not associated with a net clinical advantage across the prioritized hierarchy. The favorable signal among patients with NIHSS scores greater than 5 suggests that a clinically meaningful subgroup may exist, but this cutoff should not be treated as a stand-alone treatment rule. A patient with an NIHSS score of 4, fluctuating brainstem symptoms, poor collaterals, or a large perfusion deficit may differ substantially from a stable patient with an NIHSS score of 7, good collaterals, and a technically challenging atherosclerotic occlusion.^25^ Future treatment algorithms should integrate neurological severity with imaging and procedural predictors rather than rely on NIHSS thresholds alone.

Several limitations deserve emphasis. First, this was a secondary observational registry analysis. Stabilized inverse probability weighting improved measured covariate balance, but residual confounding by indication cannot be excluded. Second, the exposure was binary EVT versus BMM. Because prior work suggests that treatment timing may strongly influence benefit, the present analysis should not be interpreted as a direct comparison of early EVT, delayed EVT, and BMM. Door-to-puncture time was defined only among EVT-treated patients and therefore could not be incorporated as a conventional baseline treatment-selection covariate. Third, exact death dates were unavailable. Death status was ranked by observed intervals, namely death within 90 days, death after 90 days and within 1 year, and alive at 1 year. This approach preserves the priority of survival but is less granular than a true time-to-death hierarchy. Fourth, the subgroup analysis by NIHSS score was exploratory. Although the direction of effect was clinically coherent, it should not be interpreted as definitive evidence of treatment-effect modification without prospective validation. Fifth, the WR depends on the selected hierarchy and on the granularity and reliability of its components. Sensitivity analyses using alternative endpoint orders supported the main interpretation, but other clinically justified hierarchies, such as those emphasizing disabling stroke, recurrent ischemic events, or longer-term quality of life, might yield different estimates. Sixth, the analysis could not fully evaluate technical features of EVT, including reperfusion grade, first-pass effect, rescue therapy, anesthesia strategy, number of passes, or periprocedural antithrombotic management. Frontline thrombectomy strategy can affect complete revascularization, underscoring the potential importance of procedural heterogeneity in posterior-circulation occlusion.^26^ Finally, the NIHSS underweights symptoms such as truncal ataxia and dysphagia, which may cause misclassification of posterior-circulation severity and influence prognosis.

Overall, EVT was not associated with a net clinical advantage over BMM across all patients with acute VBAO and admission NIHSS scores of 0-10. The exploratory signal among patients with NIHSS scores of 6-10 suggests that EVT may be feasible in selected patients, but randomized trials are needed. For patients with NIHSS scores of 0-5, EVT use should remain cautious and individualized. Future prospective studies should combine neurological severity with imaging risk, stroke mechanism, treatment timing, and reperfusion quality to identify patients in whom EVT provides a true net benefit.

### Sources of Funding

This study was funded by the Anhui Province Clinical Medicine Transformation Special Project in China (No. 202427b10020048), Key Project of the Department of Education of Anhui Province (No.2025AHGXZK20227), Guangzhou Municipal Science and Technology Bureau Key Research and Development Program (2024B03J0436) and the Research Funds of Centre for Leading Medicine and Advanced Technologies of IHM in China (No.2023IHM01053).

## Acknowledgments

Yapeng Guo and Xinyu Fan contributed to the study conception and design, methodology, formal analysis, visualization, and writing of the original draft. Yingjie Xu, Xinru Zhou, Wei Li, Junfeng Xu, and Zhixin Huang contributed to investigation, data curation, validation, and critical revision of the manuscript. Wensheng Zhang and Xianjun Huang contributed to the study conception, supervision, project administration, and critical revision of the manuscript. All authors interpreted the data, reviewed and approved the final manuscript, and agree to be accountable for all aspects of the work.

## Data Availability Statement

The data used in this secondary analysis are de-identified registry data governed by the parent study and the participating institutions. The data are not publicly available because of institutional and ethical restrictions. Subject to the approvals and data-sharing policies of the parent registry and participating centers, de-identified data may be available from the corresponding author on reasonable request.

## Ethics Statement and Consent

The parent registry was approved by the ethics committee of the First Affiliated Hospital of the University of Science and Technology of China and participating centers (No. 2020-40). The requirement for informed consent was waived because of the retrospective study design.

## Nonstandard Abbreviations and Acronyms

BMM: best medical management
EVT: endovascular thrombectomy
IPTW: inverse probability of treatment weighting
mRS: modified Rankin Scale
NIHSS: National Institutes of Health Stroke Scale
sICH: symptomatic intracranial hemorrhage
VBAO: vertebrobasilar artery occlusion
WO: win odds
WR: win ratio

